# CD8+ T cell responses in COVID-19 convalescent individuals target conserved epitopes from multiple prominent SARS-CoV-2 circulating variants

**DOI:** 10.1101/2021.02.11.21251585

**Authors:** Andrew D Redd, Alessandra Nardin, Hassen Kared, Evan M Bloch, Andrew Pekosz, Oliver Laeyendecker, Brian Abel, Michael Fehlings, Thomas C Quinn, Aaron AR Tobian

**Author notes:** Corresponding author: Andrew D. Redd, Rangos 540, 851 N Wolfe St, Baltimore MD, 21205. Alternative contact, Aaron AR Tobian, Rangos 540, 851 N Wolfe St, Baltimore MD 21205.

## Abstract

This study examined whether CD8+ T-cell responses from COVID-19 convalescent individuals(n=30) potentially maintain recognition of the major SARS-CoV-2 variants. Out of 45 mutations assessed, only one from the B.1.351 Spike overlapped with a low-prevalence CD8+ epitope, suggesting that virtually all anti-SARS-CoV-2 CD8+ T-cell responses should recognize these newly described variants.

## Introduction

Due to the proofreading ability of the coronavirus (CoV) RNA-dependent RNA polymerase, the evolution of the global SARS-CoV-2 viral population during the current pandemic has been relatively constrained as compared to other endemic RNA viruses that do not possess this ability[1]. However, during late 2020, three distinct variants that each possessed a significantly increased amount of amino acid polymorphisms were identified in association with spikes in cases of COVID-19 in the United Kingdom (variant B.1.1.7), South Africa (variant B.1.351), and Brazil (variant B.1.1.248)[2–4].

These variants all possess the N501Y mutation in the receptor-binding domain (RBD) of the SARS-CoV-2 spike protein, a primary target for neutralizing antibody (NAb) binding. They also all contain unique additional mutations throughout the genome, and are not phylogenetically linked, indicating that they evolved independently (Table S1). Due in part to the mutations found in the RBD and other areas of the spike protein, these primary viral variants, or pseudoviruses expressing the combination of polymorphisms found in each variant, have been examined for their sensitivity to NAb responses detected in plasmas from COVID-19 convalescent donors, preclinical or clinical trial post-vaccination plasmas, and monoclonal antibodies[4–7]. These studies have shown that the variants are variably susceptible to neutralization, with B.1.1.7 exhibiting only minor decreases in susceptibility to convalescent and post-vaccination plasma. In addition, preliminary press reports from ongoing Phase 2b/3 vaccine trials performed in the United Kingdom during the rise of B.1.1.7 suggest that the efficacy of spike-based vaccines has not diminished significantly; however, these studies have not been published or peer-reviewed. Conversely, the B.1.351 variant has demonstrated a significant increase in resistance to neutralization by some individuals’ convalescent plasma, as well as a decline in neutralization potential for the mRNA based vaccine induced NAb responses; although it should be noted that this neutralization potential was still estimated to be high enough to confer complete protection[4–7]. Of greater concern, two ongoing Phase 2/3 vaccine trials of a recombinant protein-based and an Adenovirus-based vaccine have reported a slight decrease in efficacy of the vaccines in preventing symptomatic COVID-19 in South Africa where the infections were predominantly caused by B.1.351. These data have not been published or independently reviewed. Interestingly, these same reports claimed that there was no difference in the effectiveness of the vaccines to prevent serious illness between the two countries, suggesting that while protection from initial infection may be somewhat hindered, the vaccines ability to prevent further disease progression is preserved.

While the correlates of protection in convalescent individuals and vaccinees are unknown, it is assumed that both a broad humoral and cell-mediated immunological response are most likely necessary to fully protect against COVID-19. NAb almost certainly serves as the first line of defense against infection, but the CD8+ T cell response is also important for prevention of further disease progression.

Previously, our group reported a detailed analysis of the CD8+ T cell epitopes and cytokine responses to the original strain of SARS-CoV-2 in a collection of convalescent individuals from North America with varying levels of disease and NAb responses[8]. This earlier report identified a broad CD8+ T cell response in these individuals with virtually all subjects having detectable responses to several viral epitopes.

## Methods

Detailed methods of the previous study have been previously published[8]. Briefly, peripheral blood mononuclear cell (PBMC) samples from PCR-confirmed recovered COVID-19 convalescent individuals (n=30) were collected and examined across six different HLA haplotypes (HLA-A*01:01, HLA-A*02:01, HLA-A03:01, HLA-A*11:01, HLA-A*24:02 and HLA-B*07:02). A multiplexed peptide-MHC tetramer staining approach permitted the screening of 408 potential SARS-CoV-2 candidate epitopes for CD8+ T cell recognition. T cells were also evaluated using a 28-marker phenotypic panel. 52 unique epitope responses were found and were directed against several structural and non-structural viral proteins. For controls, CD8+ T cells were probed for reactivity for up to 20 different SARS-CoV-2-unrelated control peptides per HLA (Adenovirus-, CMV-, EBV-, Influenza-, and MART-1-derived epitopes).

Amino acid polymorphisms found in the B.1.1.7, B.1.351, and B.1.1.248 variants were mapped to the SARS-CoV-2 genome. The variants were then examined for overlap with the identified epitopes from the previous study (Figure S1; Table S1).

This study was approved by the Johns Hopkins Institutional Review Board, and all participants provided informed consent.

## Results

As previously described, 60% of individuals included in the analysis were male and samples were collected a median of 42.5 days (interquartile range 37.5-48.0) from their initial diagnosis[8]. The group was selected evenly from tertiles (10 each) according to their overall anti-SARS-CoV-2 IgG titers[8,9]. Among the convalescent individuals, there were 132 SARS-CoV-2-specific CD8+ T cell responses corresponding to 52 unique epitope reactivities directed against several structural and non-structural target epitopes from the entire proteome.

Of all the mapped mutations, insertions, and deletions (n=45), only one mutation was found to fall within one of the 52 unique epitopes identified in the previous study (Fig 1, S1). This mutation is the D80A mutation in the viral Spike protein, and occurs in the third residue of the RFDNPVLPF epitope. This is a HLA*A24:02-restricted epitope for which a CD8+ T cell response was detected in only one individual, and at a low frequency, indicating this is not a high-prevalence epitope within the studied cross-sectional sample.

**Figure 1.**
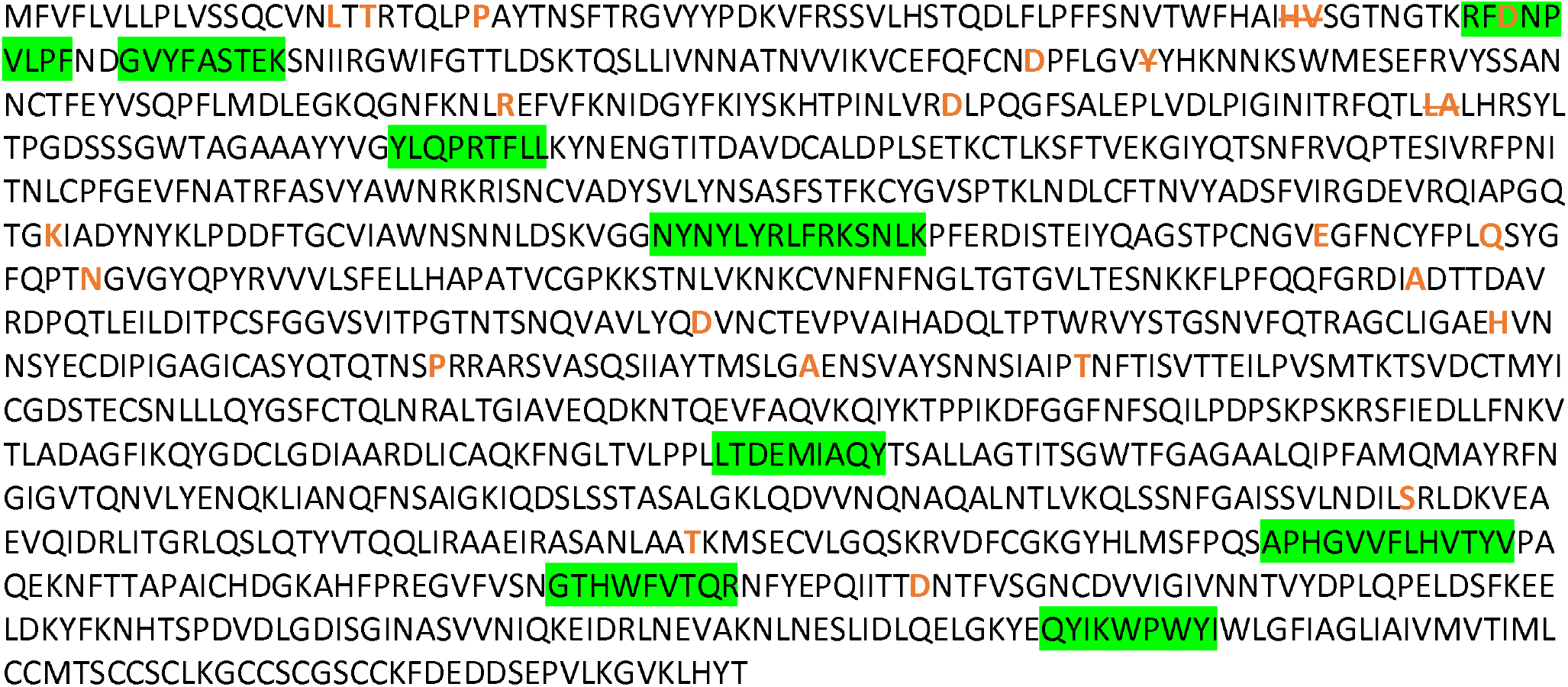
SARS-CoV-2 Wuhan variant Spike protein amino acid sequence with CD8+ T cell epitopes highlighted in green, and all mutation and deletion sites (slash) indicated in bold orange.

## Discussion

As the global SARS-CoV-2 pandemic continues, it is inevitable that new viral variants will emerge. Many of these variants will disappear unnoticed due to a combination of incomplete or non-existent genotypic surveillance, changes that have deleterious impact on viral fitness, and conditions that do not promote their further spread in the population. However, some variants, like the three examined here, will emerge in situations where continued spread is possible due to either differences in underlying infectivity of the variant, large founder effects, incomplete treatment interventions, immunocompromised patient groups, low levels of societal physical interventions, or a combination of all or some of these factors. Understanding the extant immunity in previously infected individuals to any new variant is of critical importance to properly estimate what effect these variants may have in the global pandemic[10].

This analysis identified only one mutation from the three most prominent new global SARS-CoV-2 variants that overlapped with 52 CD8+ T cell epitopes previously identified in a group of convalescent individuals. Additionally, this mutation was found on the third residue of the epitope, and given that the predicted anchor residues for HLA binding of this epitope are residues two and nine, one could speculate that this mutation may not significantly affect HLA binding and subsequent TCR recognition[11].

The data on CD8+ T cell responses from these 30 convalescent individuals are also in line with another recent report showing that the great majority of CD4+ T cell epitopes from the spike protein of the SARS-CoV-2 B.1.17 variant are also conserved[12]. Yet it should be noted, that new variants are continuing to be identified all over the world, and it will be important to continually examine these for the possible accumulation of T cell escape mutations.

The preliminary analyses of NAb responses to the B.1.1.7, B.1.351, and B.1.1.248 variants have yielded varied outcomes with some reduced efficacy of monoclonal antibody therapeutics, and a significant decrease in the neutralization capacity of plasma from convalescent and vaccinated individuals to the B.1.351 variant. The vaccines that have been tested thus far against the B.1.351 variant in South Africa demonstrate reduced protective efficacy against COVID-19, but do seem to reduce the number of cases of severe disease at an equivalent level as they do against other strains[4–7]. Yet, it must be highlighted many of these results have not been published in full detail or peer-reviewed. This possible prevention of severe disease may be due in part to the cell-mediated immunity generated due to natural COVID-19 infection or vaccination, which the results presented here would suggest are minimally affected by the mutations found in these variants.

It should be noted that this study had several limitations including the relatively small size of the population examined. In addition, the participants in the study were all from North America and were selected in part on the presence of one or more of the target HLA types examined (73% coverage of the continental US population). It will be important to examine for T cell escape in more diverse HLA types moving forward.

These data highlight the potential significant role of a multi-epitope T cell response in limiting viral escape, and partly mediate protection from disease caused by the SARS-CoV-2 variants. It is important that vaccines used for widespread campaigns generate strong multivalent T-cell responses in addition to NAb and other humoral responses in order to optimize efficacy against the current SARS-CoV-2 and emerging strains. It will also be important to continue to monitor the breadth, magnitude, and durability of the anti-SARS-CoV-2 T cell responses in recovered and vaccinated individuals as part of any assessment to determine if booster vaccinations are needed.

## Data Availability

The data used in this analysis is available in a previous publication
"SARS-CoV-2-specific CD8+ T cell responses in convalescent COVID-19 individuals"

https://www.jci.org/articles/view/145476

## Acknowledgements

The authors would like to thank the CCP study and laboratory teams, and all the donors for the generous participation in the study.

