## Supplemental Table for "CD8+ T cell responses in COVID-19 convalescent individuals target conserved epitopes from multiple prominent SARS-CoV-2 circulating variants"

| **Gene** | **Type** | **Amino acid** | **Variant** | **Source** |
| --- | --- | --- | --- | --- |
| **N** |  | P80R | Brazil | https://virological.org/t/genomic-characterisation-of-an-emergent-sars-cov-2-lineage-in-manaus-preliminary-findings/586 |
| **ORF1ab** | Deletion | SGF 3675–3677 deletion | Brazil | https://virological.org/t/genomic-characterisation-of-an-emergent-sars-cov-2-lineage-in-manaus-preliminary-findings/586 |
| **ORF1ab** |  | E5665D | Brazil | https://virological.org/t/genomic-characterisation-of-an-emergent-sars-cov-2-lineage-in-manaus-preliminary-findings/586 |
| **ORF1ab** |  | K1795Q | Brazil | https://virological.org/t/genomic-characterisation-of-an-emergent-sars-cov-2-lineage-in-manaus-preliminary-findings/586 |
| **ORF1ab** |  | S1188L | Brazil | https://virological.org/t/genomic-characterisation-of-an-emergent-sars-cov-2-lineage-in-manaus-preliminary-findings/586 |
| **Orf3a** |  | G174C | Brazil | https://virological.org/t/genomic-characterisation-of-an-emergent-sars-cov-2-lineage-in-manaus-preliminary-findings/586 |
| **Orf8** | Insertion |  | Brazil | https://virological.org/t/genomic-characterisation-of-an-emergent-sars-cov-2-lineage-in-manaus-preliminary-findings/586 |
| **Orf8** |  | E92K | Brazil | https://virological.org/t/genomic-characterisation-of-an-emergent-sars-cov-2-lineage-in-manaus-preliminary-findings/586 |
| **Spike** |  | D138Y | Brazil | https://virological.org/t/genomic-characterisation-of-an-emergent-sars-cov-2-lineage-in-manaus-preliminary-findings/586 |
| **Spike** |  | E484K | Brazil | https://virological.org/t/genomic-characterisation-of-an-emergent-sars-cov-2-lineage-in-manaus-preliminary-findings/586 |
| **Spike** |  | H655Y | Brazil | https://virological.org/t/genomic-characterisation-of-an-emergent-sars-cov-2-lineage-in-manaus-preliminary-findings/586 |
| **Spike** |  | K417T | Brazil | https://virological.org/t/genomic-characterisation-of-an-emergent-sars-cov-2-lineage-in-manaus-preliminary-findings/586 |
| **Spike** |  | L18F | Brazil | https://virological.org/t/genomic-characterisation-of-an-emergent-sars-cov-2-lineage-in-manaus-preliminary-findings/586 |
| **Spike** |  | N501Y | Brazil | https://virological.org/t/genomic-characterisation-of-an-emergent-sars-cov-2-lineage-in-manaus-preliminary-findings/586 |
| **Spike** |  | P26S | Brazil | https://virological.org/t/genomic-characterisation-of-an-emergent-sars-cov-2-lineage-in-manaus-preliminary-findings/586 |
| **Spike** |  | R190S | Brazil | https://virological.org/t/genomic-characterisation-of-an-emergent-sars-cov-2-lineage-in-manaus-preliminary-findings/586 |
| **Spike** |  | T1027I | Brazil | https://virological.org/t/genomic-characterisation-of-an-emergent-sars-cov-2-lineage-in-manaus-preliminary-findings/586 |
| **Spike** |  | T20N | Brazil | https://virological.org/t/genomic-characterisation-of-an-emergent-sars-cov-2-lineage-in-manaus-preliminary-findings/586 |
| **E** |  | P71L | SA | https://www.ahri.org/wp-content/uploads/2021/01/MEDRXIV-2021-250224v1-Sigal.pdf |
| **N** |  | T205I | SA | https://www.ahri.org/wp-content/uploads/2021/01/MEDRXIV-2021-250224v1-Sigal.pdf |
| **ORF14** |  | L52F | SA | https://www.ahri.org/wp-content/uploads/2021/01/MEDRXIV-2021-250224v1-Sigal.pdf |
| **ORF1a** |  | K1655N | SA | https://www.ahri.org/wp-content/uploads/2021/01/MEDRXIV-2021-250224v1-Sigal.pdf |
| **ORF1a** |  | K3353R | SA | https://www.ahri.org/wp-content/uploads/2021/01/MEDRXIV-2021-250224v1-Sigal.pdf |
| **ORF1a** |  | T265I | SA | https://www.ahri.org/wp-content/uploads/2021/01/MEDRXIV-2021-250224v1-Sigal.pdf |
| **ORF1ab** | Deletion | SGF 3675–3677 deletion | SA | https://www.ahri.org/wp-content/uploads/2021/01/MEDRXIV-2021-250224v1-Sigal.pdf |
| **ORF1b** |  | P314L | SA | https://www.ahri.org/wp-content/uploads/2021/01/MEDRXIV-2021-250224v1-Sigal.pdf |
| **ORF3a** |  | Q57H | SA | https://www.ahri.org/wp-content/uploads/2021/01/MEDRXIV-2021-250224v1-Sigal.pdf |
| **ORF3a** |  | S171L | SA | https://www.ahri.org/wp-content/uploads/2021/01/MEDRXIV-2021-250224v1-Sigal.pdf |
| **ORF3a** |  | W131L | SA | https://www.ahri.org/wp-content/uploads/2021/01/MEDRXIV-2021-250224v1-Sigal.pdf |
| **ORF7a** |  | V93F | SA | https://www.ahri.org/wp-content/uploads/2021/01/MEDRXIV-2021-250224v1-Sigal.pdf |
| **Spike** | Deletion | 242-244 deletion | SA | https://www.ahri.org/wp-content/uploads/2021/01/MEDRXIV-2021-250224v1-Sigal.pdf |
| **Spike** | Deletion | 69-70 deletion | SA | https://www.ahri.org/wp-content/uploads/2021/01/MEDRXIV-2021-250224v1-Sigal.pdf |
| **Spike** |  | A701V | SA | https://www.ahri.org/wp-content/uploads/2021/01/MEDRXIV-2021-250224v1-Sigal.pdf |
| **Spike** |  | D215G | SA | https://www.ahri.org/wp-content/uploads/2021/01/MEDRXIV-2021-250224v1-Sigal.pdf |
| **Spike** |  | D614G | SA | https://www.ahri.org/wp-content/uploads/2021/01/MEDRXIV-2021-250224v1-Sigal.pdf |
| **Spike** |  | D80A | SA | https://www.ahri.org/wp-content/uploads/2021/01/MEDRXIV-2021-250224v1-Sigal.pdf |
| **Spike** |  | E484K | SA | https://www.ahri.org/wp-content/uploads/2021/01/MEDRXIV-2021-250224v1-Sigal.pdf |
| **Spike** |  | K417N | SA | https://www.ahri.org/wp-content/uploads/2021/01/MEDRXIV-2021-250224v1-Sigal.pdf |
| **Spike** |  | L18F | SA | https://www.ahri.org/wp-content/uploads/2021/01/MEDRXIV-2021-250224v1-Sigal.pdf |
| **Spike** |  | N501Y | SA | https://www.ahri.org/wp-content/uploads/2021/01/MEDRXIV-2021-250224v1-Sigal.pdf |
| **N** |  | D3L | UK | https://assets.publishing.service.gov.uk/government/uploads/system/uploads/attachment_data/file/947048/Technical_Briefing_VOC_SH_NJL2_SH2.pdf |
| **N** |  | S235F | UK | https://assets.publishing.service.gov.uk/government/uploads/system/uploads/attachment_data/file/947048/Technical_Briefing_VOC_SH_NJL2_SH2.pdf |
| **ORF10** |  | Y73C | UK | https://assets.publishing.service.gov.uk/government/uploads/system/uploads/attachment_data/file/947048/Technical_Briefing_VOC_SH_NJL2_SH2.pdf |
| **Orf1ab** | Deletion | SGF 3675–3677 deletion | UK | https://assets.publishing.service.gov.uk/government/uploads/system/uploads/attachment_data/file/947048/Technical_Briefing_VOC_SH_NJL2_SH2.pdf |
| **ORF1ab** |  | A1708D | UK | https://assets.publishing.service.gov.uk/government/uploads/system/uploads/attachment_data/file/947048/Technical_Briefing_VOC_SH_NJL2_SH2.pdf |
| **ORF1ab** |  | I2230T | UK | https://assets.publishing.service.gov.uk/government/uploads/system/uploads/attachment_data/file/947048/Technical_Briefing_VOC_SH_NJL2_SH2.pdf |
| **ORF1ab** |  | T1001I | UK | https://assets.publishing.service.gov.uk/government/uploads/system/uploads/attachment_data/file/947048/Technical_Briefing_VOC_SH_NJL2_SH2.pdf |
| **ORF8** |  | Q27stop | UK | https://assets.publishing.service.gov.uk/government/uploads/system/uploads/attachment_data/file/947048/Technical_Briefing_VOC_SH_NJL2_SH2.pdf |
| **ORF9** |  | R52I | UK | https://assets.publishing.service.gov.uk/government/uploads/system/uploads/attachment_data/file/947048/Technical_Briefing_VOC_SH_NJL2_SH2.pdf |
| **Spike** | Deletion | 69–70 deletion | UK | https://assets.publishing.service.gov.uk/government/uploads/system/uploads/attachment_data/file/947048/Technical_Briefing_VOC_SH_NJL2_SH2.pdf |
| **Spike** | Deletion | Y144 deletion | UK | https://assets.publishing.service.gov.uk/government/uploads/system/uploads/attachment_data/file/947048/Technical_Briefing_VOC_SH_NJL2_SH2.pdf |
| **Spike** |  | A570D | UK | https://assets.publishing.service.gov.uk/government/uploads/system/uploads/attachment_data/file/947048/Technical_Briefing_VOC_SH_NJL2_SH2.pdf |
| **Spike** |  | D1118H | UK | https://assets.publishing.service.gov.uk/government/uploads/system/uploads/attachment_data/file/947048/Technical_Briefing_VOC_SH_NJL2_SH2.pdf |
| **Spike** |  | N501Y | UK | https://assets.publishing.service.gov.uk/government/uploads/system/uploads/attachment_data/file/947048/Technical_Briefing_VOC_SH_NJL2_SH2.pdf |
| **Spike** |  | P681H | UK | https://assets.publishing.service.gov.uk/government/uploads/system/uploads/attachment_data/file/947048/Technical_Briefing_VOC_SH_NJL2_SH2.pdf |
| **Spike** |  | S982A | UK | https://assets.publishing.service.gov.uk/government/uploads/system/uploads/attachment_data/file/947048/Technical_Briefing_VOC_SH_NJL2_SH2.pdf |
| **Spike** |  | T716I | UK | https://assets.publishing.service.gov.uk/government/uploads/system/uploads/attachment_data/file/947048/Technical_Briefing_VOC_SH_NJL2_SH2.pdf |

**Table S1: Mutations, Insertions and deletions for three variants examined.**
