## Supplemental Figure 1 for "CD8+ T cell responses in COVID-19 convalescent individuals target conserved epitopes from multiple prominent SARS-CoV-2 circulating variants"

**Figure S1:**

Nucleoprotein

MS**D**NGPQNQRNAPRITFGGPSDSTGSNQNGERSGARSKQRRPQGLPNNTASWFTALTQHGKEDLKFPRGQGVPINTNSS**P**DDQIGYYRRATRRIRGGDGKMKDLSPRWYFYYLGTGPEAGLPYGANKDGIIWVATEGALNTPKDHIGTRNPANNAAIVLQLPQGTTLPKGFYAEGSRGGSQASSRSSSRSRNSSRNSTPGSSRG**T**SPARMAGNGGDAALALLLLDRLNQLESKM**S**GKGQQQQGQTVTKKSAAEASKKPRQKRTATKAYNVTQAFGRRGPEQTQGNFGDQELIRQGTDYKHWPQIAQFAPSASAFFGMSRIGMEVTPSGTWLTYTGAIKLDDKDPNFKDQVILLNKHIDAYKTFPPTEPKKDKKKKADETQALPQRQKKQQTVTLLPAADLDDFSKQLQQSMSSADSTQA

ORF7a

MKIILFLALITLATCELYHYQECVRGTTVLLKEPCSSGTYEGNSPFHPLADNKFALTCFSTQFAFACPDGVKHVYQLRARSVSPKLFIRQEE**V**QELYSPIFLIVAAIVFITLCFTLKRKTE

ORF3a

MDLFMRIFTIGTVTLKQGEIKDATPSDFVRATATIPIQASLPFGWLIVGVALLAVF**Q**SASKIITLKKRWQLALSKGVHFVCNLLLLFVTVYSHLLLVAAGLEAPFLYLYALVYFLQSINFVRIIMRLWLC**W**KCRSKNPLLYDANYFLCWHTNCYDYCIPYNSVTSSIVIT**S**GD**G**TTSPISEHDYQIGGYTEKWESGVKDCVVLHSYFTSDYYQLYSTQLSTDTGVEHVTFFIYNKIVDEPEEHVQIHTIDGSSGVVNPVMEPIYDEPTTTTSVPL

ORF1ab

MESLVPGFNEKTHVQLSLPVLQVRDVLVRGFGDSVEEVLSEARQHLKDGTCGLVEVEKGVLPQLEQPYVFIKRSDARTAPHGHVMVELVAELEGIQYGRSGETLGVLVPHVGEIPVAYRKVLLRKNGNKGAGGHSYGADLKSFDLGDELGTDPYEDFQENWNTKHSSGVTRELMRELNGGAYTRYVDNNFCGPDGYPLECIKDLLARAGKASCTLSEQLDFIDTKRGVYCCREHEHEIAWYTERSEKSYELQTPFEIKLAKKFD**T**FNGECPNFVFPLNSIIKTIQPRVEKKKLDGFMGRIRSVYPVASPNECNQMCLSTLMKCDHCGETSWQTGDFVKATCEFCGTENLTKEGATTCGYLPQNAVVKIYCPACHNSEVGPEHSLAEYHNESGLKTILRKGGRTIAFGGCVFSYVGCHNKCAYWVPRASANIGCNHTGVVGEGSEGLNDNLLEILQKEKVNINIVGDFKLNEEIAIILASFSASTSAFVETVKGLDYKAFKQIVESCGNFKVTKGKAKKGAWNIGEQKSILSPLYAFASEAARVVRSIFSRTLETAQNSVRVLQKAAITILDGISQYSLRLIDAMMFTSDLATNNLVVMAYITGGVVQLTSQWLTNIFGTVYEKLKPVLDWLEEKFKEGVEFLRDGWEIVKFISTCACEIVGGQIVTCAKEIKESVQTFFKLVNKFLALCADSIIIGGAKLKALNLGETFVTHSKGLYRKCVKSREETGLLMPLKAPKEIIFLEGETLPTEVLTEEVVLKTGDLQPLEQPTSEAVEAPLVGTPVCINGLMLLEIKDTEKYCALAPNMMVTNNTFTLKGGAPTKVTFGDDTVIEVQGYKSVNITFELDERIDKVLNEKCSAYTVELGTEVNEFACVVADAVIKTLQPVSELLTPLGIDLDEWSMATYYLFDESGEFKLASHMYCSFYPPDEDEEEGDCEEEEFEPSTQYEYGTEDDYQGKPLEFGATSAALQPEEEQEEDWLDDDSQQTVGQQDGSEDNQTT**T**IQTIVEVQPQLEMELTPVVQTIEVNSFSGYLKLTDNVYIKNADIVEEAKKVKPTVVVNAANVYLKHGGGVAGALNKATNNAMQVESDDYIATNGPLKVGGSCVLSGHNLAKHCLHVVGPNVNKGEDIQLLKSAYENFNQHEVLLAPLLSAGIFGADPIHSLRVCVDTVRTNVYLAVFDKNLYDKLV**S**SFLEMKSEKQVEQKIAEIPKEEVKPFITESKPSVEQRKQDDKKIKACVEEVTTTLEETKFLTENLLLYIDINGNLHPDSATLVSDIDITFLKKDAPYIVGDVVQEGVLTAVVIPTKKAGGTTEMLAKALRKVPTDNYITTYPGQGLNGYTVEEAKTVLKKCKSAFYILPSIISNEKQEILGTVSWNLREMLAHAEETRKLMPVCVETKAIVSTIQRKYKGIKIQEGVVDYGARFYFYTSKTTVASLINTLNDLNETLVTMPLGYVTHGLNLEEAARYMRSLKVPATVSVSSPDAVTAYNGYLTSSSKTPEEHFIETISLAGSYKDWSYSGQSTQLGIEFLKRGDKSVYYTSNPTTFHLDGEVITFDNLKTLLSLREVRTIKVFTTVDNINLHTQVVDMSMTYGQQFGPTYLDGADVTKIKPHNSHEGKTFYVLPNDDTLRVEAFEYYHTTDPSFLGRYMSALNHTK**K**WKYPQVNGLTSIKWADNNCYLATALLTLQQIELKFNPPALQDAYYRARAGEA**A**NFCALILAYCNKTVGELGDVRETMSYLFQHANLDSCKRVLNVVCKTCGQQQTTLKGVEAVMYMGTLSYEQFKKGVQIPCTCGKQAT**K**YLVQQESPFVMMSAPPAQYELKHGTFTCASEYTGNYQCGHYKHITSKETLYCIDGALLTKSSEYKGPITDVFYKENSYTTTIKPVTYKLDGVVCTEIDPKLDNYYKKDNSYFTEQPIDLVPNQPYPNASFDNFKFVCDNIKFADDLNQLTGYKKPASRELKVTFFPDLNGDVVAIDYKHYTPSFKKGAKLLHKPIVWHVNNATNKATYKPNTWCIRCLWSTKPVETSNSFDVLKSEDAQGMDNLACEDLKPVSEEVVENPTIQKDVLECNVKTTEVVGDIILKPANNSLKITEEVGHTDLMAAYVDNSSLTIKKPNELSRVLGLKTLATHGLAAVNSVPWDTIANYAKPFLNKVVSTTTNIVTRCLNRVCTNYMPYFFTLLLQLCTFTRSTNSRIKASMPTTIAKNTVKSVGKFCLEASFNYLKSPNFSKLINI**I**IWFLLLSVCLGSLIYSTAALGVLMSNLGMPSYCTGYREGYLNSTNVTIATYCTGSIPCSVCLSGLDSLDTYPSLETIQITISSFKWDLTAFGLVAEWFLAYILFTRFFYVLGLAAIMQLFFSYFAVHFISNSWLMWLIINLVQMAPISAMVRMYIFFASFYYVWKSYVHVVDGCNSSTCMMCYKRNRATRVECTTIVNGVRRSFYVYANGGKGFCKLHNWNCVNCDTFCAGSTFISDEVARDLSLQFKRPINPTDQSSYIVDSVTVKNGSIHLYFDKAGQKTYERHSLSHFVNLDNLRANNTKGSLPINVIVFDGKSKCEESSAKSASVYYSQLMCQPILLLDQALVSDVGDSAEVAVKMFDAYVNTFSSTFNVPMEKLKTLVATAEAELAKNVSLDNVLSTFISAARQGFVDSDVETKDVVECLKLSHQSDIEVTGDSCNNYMLTYNKVENMTPRDLGACIDCSARHINAQVAKSHNIALIWNVKDFMSLSEQLRKQIRSAAKKNNLPFKLTCATTRQVVNVVTTKIALKGGKIVNNWLKQLIKVTLVFLFVAAIFYLITPVHVMSKHTDFSSEIIGYKAIDGGVTRDIASTDTCFANKHADFDTWFSQRGGSYTNDKACPLIAAVITREVGFVVPGLPGTILRTTNGDFLHFLPRVFSAVGNICYTPSKLIEYTDFATSACVLAAECTIFKDASGKPVPYCYDTNVLEGSVAYESLRPDTRYVLMDGSIIQFPNTYLEGSVRVVTTFDSEYCRHGTCERSEAGVCVSTSGRWVLNNDYYRSLPGVFCGVDAVNLLTNMFTPLIQPIGALDISASIVAGGIVAIVVTCLAYYFMRFRRAFGEYSHVVAFNTLLFLMSFTVLCLTPVYSFLPGVYSVIYLYLTFYLTNDVSFLAHIQWMVMFTPLVPFWITIAYIICISTKHFYWFFSNYLKRRVVFNGVSFSTFEEAALCTFLLNKEMYLKLRSDVLLPLTQYNRYLALYNKYKYFSGAMDTTSYREAACCHLAKALNDFSNSGSDVLYQPPQTSITSAVLQSGFRKMAFPSGKVEGCMVQVTCGTTTLNGLWLDDVVYCPRHVICTSEDMLNPNYEDLLIRKSNHNFLVQAGNVQLRVIGHSMQNCVLKL**K**VDTANPKTPKYKFVRIQPGQTFSVLACYNGSPSGVYQCAMRPNFTIKGSFLNGSCGSVGFNIDYDCVSFCYMHHMELPTGVHAGTDLEGNFYGPFVDRQTAQAAGTDTTITVNVLAWLYAAVINGDRWFLNRFTTTLNDFNLVAMKYNYEPLTQDHVDILGPLSAQTGIAVLDMCASLKELLQNGMNGRTILGSALLEDEFTPFDVVRQCSGVTFQSAVKRTIKGTHHWLLLTILTSLLVLVQSTQWSLFFFLYENAFLPFAMGIIAMSAFAMMFVKHKHAFLCLFLLPSLATVAYFNMVYMPASWVMRIMTWLDMVDTSL**~~SGF~~**KLKDCVMYASAVVLLILMTARTVYDDGARRVWTLMNVLTLVYKVYYGNALDQAISMWALIISVTSNYSGVVTTVMFLARGIVFMCVEYCPIFFITGNTLQCIMLVYCFLGYFCTCYFGLFCLLNRYFRLTLGVYDYLVSTQEFRYMNSQGLLPPKNSIDAFKLNIKLLGVGGKPCIKVATVQSKMSDVKCTSVVLLSVLQQLRVESSSKLWAQCVQLHNDILLAKDTTEAFEKMVSLLSVLLSMQGAVDINKLCEEMLDNRATLQAIASEFSSLPSYAAFATAQEAYEQAVANGDSEVVLKKLKKSLNVAKSEFDRDAAMQRKLEKMADQAMTQMYKQARSEDKRAKVTSAMQTMLFTMLRKLDNDALNNIINNARDGCVPLNIIPLTTAAKLMVVIPDYNTYKNTCDGTTFTYASALWEIQQVVDADSKIVQLSEISMDNSPNLAWPLIVTALRANSAVKLQNNELSPVALRQMSCAAGTTQTACTDDNALAYYNTTKGGRFVLALLSDLQDLKWARFPKSDGTGTIYTELEPPCRFVTDTPKGPKVKYLYFIKGLNNLNRGMVLGSLAATVRLQAGNATEVPANSTVLSFCAFAVDAAKAYKDYLASGGQPITNCVKMLCTHTGTGQAITVTPEANMDQESFGGASCCLYCRCHIDHPNPKGFCDLKGKYVQIPTTCANDPVGFTLKNTVCTVCGMWKGYGCSCDQLREPMLQSADAQSFLNRVCGVSAARLTPCGTGTSTDVVYRAFDIYNDKVAGFAKFLKTNCCRFQEKDEDDNLIDSYFVVKRHTFSNYQHEETIYNLLKDCPAVAKHDFFKFRIDGDMVPHISRQRLTKYTMADLVYALRHFDEGNCDTLKEILVTYNCCDDDYFNKKDWYDFVENPDILRVYANLGERVRQALLKTVQFCDAMRNAGIVGVLTLDNQDLNGNWYDFGDFIQTTPGSGVPVVDSYYSLLMPILTLTRALTAESHVDTDLTKPYIKWDLLKYDFTEERLKLFDRYFKYWDQTYHPNCVNCLDDRCILHCANFNVLFSTVFPPTSFGPLVRKIFVDGVPFVVSTGYHFRELGVVHNQDVNLHSSRLSFKELLVYAADPAMHAASGNLLLDKRTTCFSVAALTNNVAFQTVKPGNFNKDFYDFAVSKGFFKEGSSVELKHFFFAQDGNAAISDYDYYRYNLPTMCDIRQLLFVVEVVDKYFDCYDGGCINANQVIVNNLDKSAGFPFNKWGKARLYYDSMSYEDQDALFAYTKRNVIPTITQMNLKYAISAKNRARTVAGVSICSTMTNRQFHQKLLKSIAATRGATVVIGTSKFYGGWHNMLKTVYSDVENPHLMGWDYPKCDRAMPNMLRIMASLVLARKHTTCCSLSHRFYRLANECAQVLSEMVMCGGSLYVKPGGTSSGDATTAYANSVFNICQAVTANVNALLSTDGNKIADKYVRNLQHRLYECLYRNRDVDTDFVNEFYAYLRKHFSMMILSDDAVVCFNSTYASQGLVASIKNFKSVLYYQNNVFMSEAKCWTETDLTKGPHEFCSQHTMLVKQGDDYVYLPYPDPSRILGAGCFVDDIVKTDGTLMIERFVSLAIDAYPLTKHPNQEYADVFHLYLQYIRKLHDELTGHMLDMYSVMLTNDNTSRYWEPEFYEAMYTPHTVLQAVGACVLCNSQTSLRCGACIRRPFLCCKCCYDHVISTSHKLVLSVNPYVCNAPGCDVTDVTQLYLGGMSYYCKSHKPPISFPLCANGQVFGLYKNTCVGSDNVTDFNAIATCDWTNAGDYILANTCTERLKLFAAETLKATEETFKLSYGIATVREVLSDRELHLSWEVGKPRPPLNRNYVFTGYRVTKNSKVQIGEYTFEKGDYGDAVVYRGTTTYKLNVGDYFVLTSHTVMPLSAPTLVPQEHYVRITGLYPTLNISDEFSSNVANYQKVGMQKYSTLQGPPGTGKSHFAIGLALYYPSARIVYTACSHAAVDALCEKALKYLPIDKCSRIIPARARV**E**CFDKFKVNSTLEQYVFCTVNALPETTADIVVFDEISMATNYDLSVVNARLRAKHYVYIGDPAQLPAPRTLLTKGTLEPEYFNSVCRLMKTIGPDMFLGTCRRCPAEIVDTVSALVYDNKLKAHKDKSAQCFKMFYKGVITHDVSSAINRPQIGVVREFLTRNPAWRKAVFISPYNSQNAVASKILGLPTQTVDSSQGSEYDYVIFTQTTETAHSCNVNRFNVAITRAKVGILCIMSDRDLYDKLQFTSLEIPRRNVATLQAENVTGLFKDCSKVITGLHPTQAPTHLSVDTKFKTEGLCVDIPGIPKDMTYRRLISMMGFKMNYQVNGYPNMFITREEAIRHVRAWIGFDVEGCHATREAVGTNLPLQLGFSTGVNLVAVPTGYVDTPNNTDFSRVSAKPPPGDQFKHLIPLMYKGLPWNVVRIKIVQMLSDTLKNLSDRVVFVLWAHGFELTSMKYFVKIGPERTCCLCDRRATCFSTASDTYACWHHSIGFDYVYNPFMIDVQQWGFTGNLQSNHDLYCQVHGNAHVASCDAIMTRCLAVHECFVKRVDWTIEYPIIGDELKINAACRKVQHMVVKAALLADKFPVLHDIGNPKAIKCVPQADVEWKFYDAQPCSDKAYKIEELFYSYATHSDKFTDGVCLFWNCNVDRYPANSIVCRFDTRVLSNLNLPGCDGGSLYVNKHAFHTPAFDKSAFVNLKQLPFFYYSDSPCESHGKQVVSDIDYVPLKSATCITRCNLGGAVCRHHANEYRLYLDAYNMMISAGFSLWVYKQFDTYNLWNTFTRLQSLENVAFNVVNKGHFDGQQGEVPVSIINNTVYTKVDGVDVELFENKTTLPVNVAFELWAKRNIKPVPEVKILNNLGVDIAANTVIWDYKRDAPAHISTIGVCSMTDIAKKPTETICAPLTVFFDGRVDGQVDLFRNARNGVLITEGSVKGLQPSVGPKQASLNGVTLIGEAVKTQFNYYKKVDGVVQQLPETYFTQSRNLQEFKPRSQMEIDFLELAMDEFIERYKLEGYAFEHIVYGDFSHSQLGGLHLLIGLAKRFKESPFELEDFIPMDSTVKNYFITDAQTGSSKCVCSVIDLLLDDFVEIIKSQDLSVVSKVVKVTIDYTEISFMLWCKDGHVETFYPKLQSSQAWQPGVAMPNLYKMQRMLLEKCDLQNYGDSATLPKGIMMNVAKYTQLCQYLNTLTLAVPYNMRVIHFGAGSDKGVAPGTAVLRQWLPTGTLLVDSDLNDFVSDADSTLIGDCATVHTANKWDLIISDMYDPKTKNVTKENDSKEGFFTYICGFIQQKLALGGSVAIKITEHSWNADLYKLMGHFAWWTAFVTNVNASSSEAFLIGCNYLGKPREQIDGYVMHANYIFWRNTNPIQLSSYSLFDMSKFPLKLRGTAVMSLKEGQINDMILSLLSKGRLIIRENNRVVISSDVLVNN

Figure S1: SARS-CoV-2 Wuhan variant protein amino acid sequence with CD8+ T cell epitopes highlighted and all mutations, insertions (box) and deletions (slash) sites indicated in bold orange (non-overlapping).
